# Automated detection of bicuspid aortic valve from echocardiographic reports using natural language processing: a large-scale Veterans Affairs study

**DOI:** 10.1101/2025.06.30.25330573

**Authors:** Annie E. Bowles, Julie A. Lynch, Francisca Bermudez, Gabrielle E. Shakt, Tia DiNatale, Kathryn M. Pridgen, Renae L. Judy, Michael G. Levin, Katherine Hartmann, Scott M. Damrauer, Patrick R. Alba

## Abstract

**Background:** Bicuspid aortic valve (BAV) is the most common congenital heart defect but often evades timely diagnosis due to variable clinical presentations. Prior to October 2024, no specific diagnosis code existed for BAV, limiting retrospective identification.

**Objectives:** To develop and validate a natural language processing (NLP) system for automated extraction of heart valve morphology from echocardiographic reports, with focus on BAV detection.

**Methods:** We developed a rule-based NLP system using MedSpaCy to analyze echocardiographic reports from the Veterans Affairs Corporate Data Warehouse. The system was trained on 555 manually annotated reports and validated on 170 held-out reports. Performance was measured using precision, recall, and F1-score for valve leaflet structure identification.

**Results:** The NLP system achieved excellent performance for BAV detection with precision of 0.984, recall of 0.955, and F1-score of 0.969. When applied to 14,453,591 echocardiographic documents from 3,478,658 patients, the system identified 84,019 patients (2.42%) with affirmed BAV. Among patients identified by the ICD-10 code Q23.81, NLP showed 86.1% concordance, with manual review confirming NLP accuracy in discordant cases.

**Conclusions:** This NLP approach enables large-scale retrospective identification of BAV patients from clinical text, creating the largest BAV cohort to date and facilitating future cardiovascular research and clinical decision-making.

## Introduction

Advances in echocardiography have substantially improved screening capabilities for valvular heart disease over the past decade. However, the unstructured free-text format of echocardiographic reports creates significant barriers to systematic data analysis and large-scale research applications.^1,2^ Lack of standardization in report formatting and time-intensive manual extraction limit our ability to analyze echocardiographic data on a larger scale.

Bicuspid aortic valve (BAV) is the most common congenital cardiac malformation, affecting 0.5-2% of the general population.^3^ This condition results from developmental fusion of two aortic valve cusps, creating a central raphe and eliminating a functional commissure. The resulting structural abnormalities disrupt normal aortic valve hemodynamics and predispose patients to serious cardiovascular complications including aortic stenosis, aortic regurgitation, and thoracic aortic aneurysm formation.^4^ Given the wide spectrum of clinical presentations—with many patients remaining asymptomatic for years—BAV frequently escapes early detection,^5,6^ emphasizing the critical importance of systematic screening and identification strategies.

The primary diagnostic modality for BAV is transthoracic echocardiography, with findings typically documented in clinical reports.^4,5^ Until October 2024, the International Classification of Diseases (ICD) lacked a specific code for BAV, creating substantial challenges for retrospective patient identification and epidemiological studies. Natural language processing (NLP) has emerged as a powerful tool for extracting structured information from clinical text, with several studies successfully applying NLP to echocardiographic reports for valve assessment and quantitative measurements.^7–13^ Recent investigations have expanded beyond single-valve analysis to comprehensive evaluation of all four cardiac valves.^1,14^ These advances demonstrate the potential for large-scale automated analysis of cardiac valve data, creating new opportunities for studying complex conditions like BAV.

This study presents a novel NLP framework specifically designed to extract heart valve morphological information from echocardiographic reports, with particular emphasis on accurate BAV identification. Using a comprehensive dataset from the U.S. Department of Veterans Affairs (VA) health system, we developed and rigorously validated an automated approach for systematic extraction of BAV-related clinical information. We examine system performance characteristics, implementation challenges, and the broader implications for improving BAV diagnosis and cardiovascular research.

## Methods

### Study Setting and Data Source

The VA operates the largest integrated healthcare system in the United States, encompassing 170 medical centers and 1,380 outpatient clinics across the United States, territories, and Philippines. The Corporate Data Warehouse (CDW) maintains comprehensive electronic health record (EHR) data for over 25 million patients dating back to 1994. This retrospective study was conducted using VA CDW clinical data with institutional review board approval and waiver of informed consent and HIPAA authorization from the Philadelphia and Salt Lake City VA Medical Centers.

### Study Population and Data Selection

The study cohort included all patients who underwent at least one echocardiography procedure within the VA healthcare system. Procedures were identified using a comprehensive list of Current Procedural Terminology (CPT) codes (see Appendix). Associated echocardiographic reports were extracted from CDW. Given the primary research focus on BAV identification, an additional keyword search was implemented to capture documents containing variants of “bicuspid” and “bileaflet” terminology (complete keyword list provided in Appendix).

The final cohort comprised 14,453,591 echocardiographic documents, corresponding to 3,478,658 unique patients. For model development and validation, 850 reports were randomly selected for manual annotation using a stratified approach: 555 reports (65%) for training, 125 reports (15%) for validation, and 170 reports (20%) for final testing. Patient-level separation was maintained between training and testing cohorts to ensure unbiased performance evaluation.

As additional validation, we identified VA patients who received the newly implemented BAV-specific ICD-10 code (Q23.81) following echocardiographic procedures and compared these cases with NLP-identified BAV patients.

### Manual Annotation Protocol

Clinical reports were annotated using the eHOST annotation platform^15^ by two registered nurses with extensive chart review experience. The annotation guideline specified identification of four cardiac valves (aortic, mitral, tricuspid, pulmonary), associated leaflet structures when present, and creation of relationships between valve and structure entities. Given the limited terminology used for cardiac valve and leaflet structure description, the eHOST pre-annotation tool was utilized to highlight known keywords prior to manual review, with annotators instructed to identify any missed entities.

Context statements including uncertainty, negation, historical mentions, or non-patient experiencers were captured and linked to corresponding valve entities. Prosthetic heart valves were annotated as distinct entities. For bicuspid leaflet structures, annotators specified whether the valve was functionally or congenitally bicuspid (default: unspecified). The final annotation schema included 14 entities, 2 relationship types (valve-to-leaflet and valve-to-context), and 1 attribute classification. A total of 1,641 annotations were created, with 315 utilized in the held-out test dataset.

### NLP System Architecture

The NLP system was designed as a rule-based framework for identifying cardiac valves and associated leaflet morphology in echocardiographic reports. Development utilized MedSpaCy,^16^ a Python clinical NLP library built on the spaCy framework. **Figure 1** illustrates the system architecture and processing logic.

**Figure 1.**
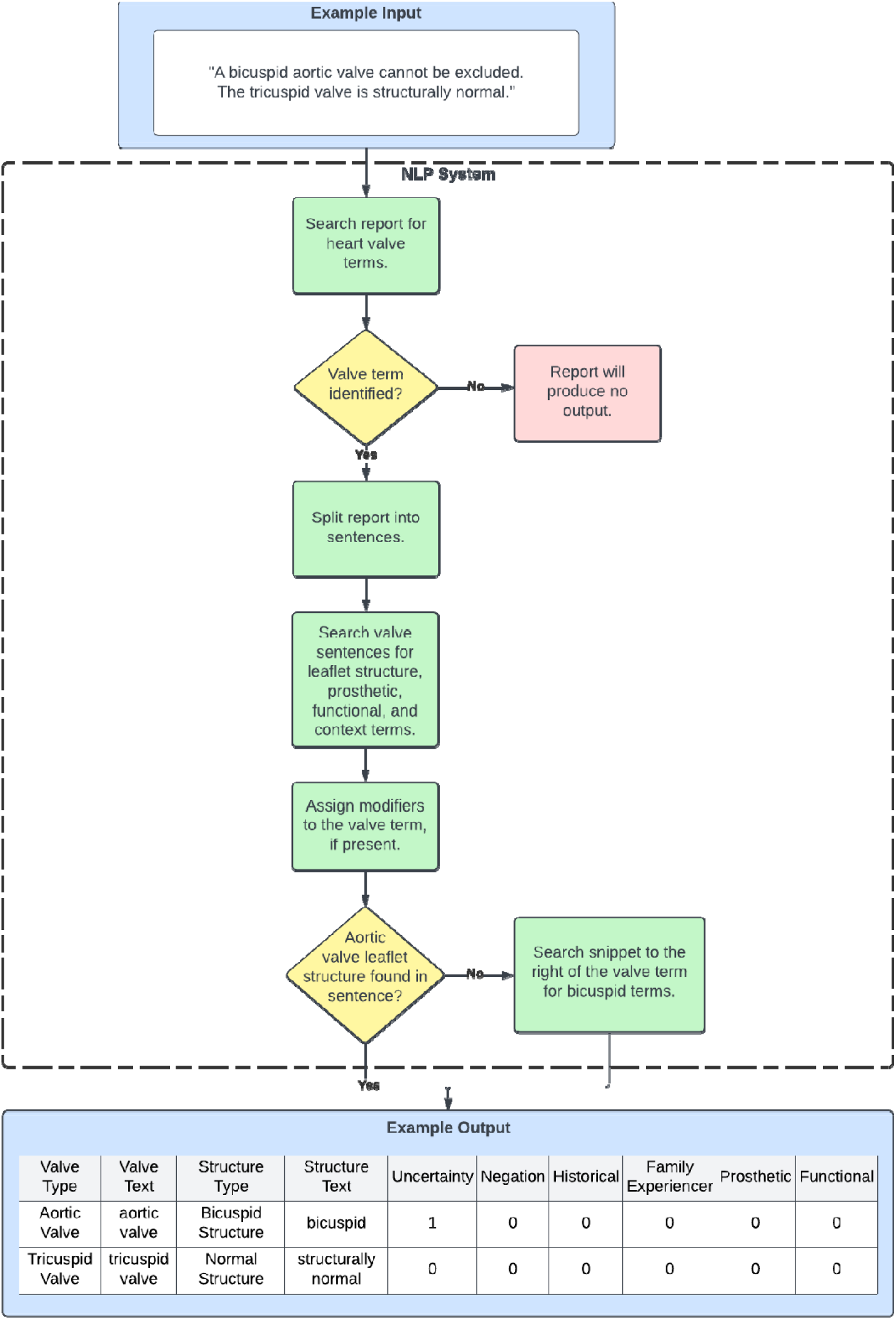
Overview of the NLP system logic with an example of the input and output structure.

The system initiates document processing by identifying mentions of the four cardiac valves: aortic, tricuspid, pulmonary, and mitral. While valve acronyms were accepted, complete valve phrases were required for entity recognition (isolated terms like “aortic” were excluded). Each identified heart valve served as an anchor point for subsequent concept attachment, with valve identification representing the minimum system output.

Following valve identification, the system searched the containing sentence for leaflet structure terms: bicuspid, tricuspid, and normal. Priority assignment favored more specific terms (bicuspid, tricuspid) based on proximity to the valve mention. Normal structure classification was assigned only when specific terms were absent. To distinguish functional BAV from congenital variants, the system searched for terms indicating functional bicuspid morphology. Prosthetic valve identification was implemented to differentiate mechanical from native BAV, as many prosthetic aortic valves exhibit bicuspid structure.

The ConText algorithm^17^ was then applied at the sentence level to identify uncertainty, negation, historical, and non-patient experiencer modifiers related to valve leaflet structure. To enhance BAV detection sensitivity, a custom component analyzed aortic valve instances lacking sentence-level leaflet structure assignment by creating a 100-token window extension and flagging bicuspid terminology within this expanded context.

System output included the document identification, valve type and raw text, leaflet structure type and raw text, context flags, functional, and prosthetic indicators, the sentence containing the heart valve, and the heart valve start and end indices. The NLP system has been made publicly available at https://github.com/VINCI-AppliedNLP/bicuspid-aortic-valve.

## Results

### NLP System Performance

The system was evaluated on the 315 instances from the held-out test set where both heart valve and leaflet structure annotations were present. **Table 1** presents detailed performance metrics at the instance level, stratified by valve type. Given the primary focus on BAV identification, the bicuspid aortic valve subset is presented separately within the aortic valve group.

**Table 1.**
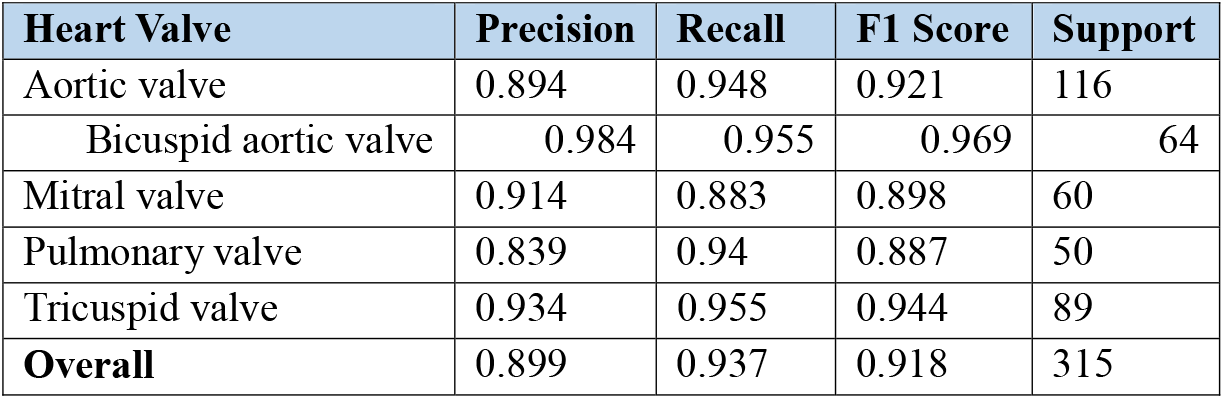
System performance on the identification of each heart valve and corresponding leaflet structure. Support is the number of annotated heart valves where the leaflet structure is given.

For BAV detection specifically, the system achieved recall of 0.955 and precision of 0.984, corresponding to an F1-score of 0.969. Overall system performance across all valve types demonstrated precision of 0.899, recall of 0.937, and F1-score of 0.918. The total support count of 315 represents non-overlapping instances, with bicuspid aortic valve cases constituting a subset of the broader aortic valve category.

### Patient-Level BAV Classification

Application of the NLP system to the complete dataset of 14,453,591 documents identified 655,762 documents (4.54%) and 84,019 patients (2.42%) with affirmed BAV. Patient-level BAV classification required at least one instance of aortic valve with bicuspid structure that was neither prosthetic nor functionally bicuspid. Given the congenital nature of BAV, instances with historical modifiers were considered affirmed cases. Patients with exclusively uncertain BAV mentions were classified as possible BAV. For patients who underwent aortic valve replacement procedures, affirmed BAV classification required evidence of congenital BAV in pre-procedural reports.

### Comparison with ICD-10 BAV Classification

Analysis of patients assigned the BAV-specific ICD-10 code (Q23.81) identified 1,573 individuals who received this diagnosis following echocardiographic procedures. Among these, 1,355 patients (86.1%) were concordantly identified as BAV cases by the NLP system.

To investigate the 218 discordant cases (ICD-10 positive, NLP negative), 50 patients were randomly selected for detailed manual review of corresponding echocardiographic reports and NLP output. The majority of these reports explicitly documented tricuspid aortic valve morphology or structurally normal valves, with the NLP system correctly extracting this information. Less frequently, reports indicated poor aortic valve visualization or absent morphological description, resulting in appropriate null NLP output. Importantly, no discordant cases were attributed to NLP system errors, suggesting potential issues with ICD-10 coding accuracy or documentation of BAV diagnosis in reports not captured by our selection criteria.

## Discussion

We successfully developed and validated an NLP system capable of accurately extracting heart valve leaflet morphology from echocardiographic reports, with particular strength in BAV identification. Implementation across the VA healthcare system identified 84,019 patients with affirmed BAV, representing the largest BAV cohort assembled to date, exceeding previous studies by more than ten-fold.^18–21^ Previous BAV research has relied on CPT codes for aortic valve replacement procedures and ICD codes related to congenital cardiac malformations for patient identification.^18^ However, these approaches lack specificity for BAV and may substantially underestimate disease prevalence. Our NLP approach enables researchers to create focused BAV cohorts that extend well beyond the limitations of structured EHR data elements. The high-performance metrics achieved on the independent test set (F1-score of 0.969 for BAV) demonstrate the system’s reliability for accurate case identification.

The recent introduction of the BAV-specific ICD-10 code (Q23.81) occurred near the completion of our study, resulting in limited overlap between NLP-identified and ICD-coded patients within our dataset. Manual review of discordant cases revealed that ICD-10 positive but NLP-negative patients had clear documentation of tricuspid or normal aortic valve morphology, suggesting either documentation of BAV in external reports not captured by our analysis or potential diagnostic coding errors. While this ICD-10 code provides a new avenue for BAV identification, it only became available in October 2024. Our NLP system enables comprehensive retrospective identification of BAV patients using historical EHR data predating this coding implementation.

This work demonstrates the potential for NLP technology to unlock valuable clinical information embedded within unstructured medical text. The ability to systematically identify large BAV cohorts from historical echocardiographic data creates new opportunities for cardiovascular research, including studies of disease progression, treatment outcomes, and genetic associations. The automated approach also supports clinical decision-making by facilitating identification of patients who may benefit from specialized cardiovascular care or surveillance protocols.

### Error Analysis and System Limitations

Analysis of NLP output errors revealed that the majority involved tricuspid and normal leaflet structure classification. The dual meaning of “tricuspid” as both a valve name and structure type created classification challenges when both references appeared in the same sentence (e.g., “TRICUSPID VALVE: tricuspid is normal in morphology”). More sophisticated disambiguation logic would be required to address these instances.

Similarly, “normal” references to concepts other than leaflet structure (e.g., “Aortic valve excursion is normal”) were incorrectly classified as normal leaflet morphology. This issue particularly affected pulmonary valve classification, which had the lowest annotation support (50 instances) and frequent references to normal functional parameters. The system’s prioritization of specific terms (tricuspid, bicuspid) before general terms (normal) improved overall performance but requires refinement for comprehensive valve assessment.

Additional error sources included incomplete valve phrase identification, unrecognized prosthetic valves, and spelling variants not captured during training. These issues were relatively infrequent but highlight areas for future system enhancement.

### Study Limitations

Several limitations merit consideration. The NLP approach only identifies BAV in patients who underwent echocardiographic procedures within the VA system, potentially missing cases diagnosed through external healthcare providers. The system was specifically developed and validated using echocardiographic reports, and performance may decrease when applied to other clinical note types.

To minimize overall errors, the system assigns each structure term to only one valve entity, though some reports use single structure terms to describe multiple valves (e.g., “AV, MV, TV, PV are normal”). More sophisticated logic would be needed to handle these instances, though their rarity in our dataset did not justify implementation complexity.

The literature describes unicuspid and quadricuspid valve morphologies as extremely rare variants. No instances were identified during development, so these structure types are not included in the current system. The prosthetic valve detection rules were optimized for aortic valves given the BAV focus, and additional development would be needed for comprehensive prosthetic valve identification across all cardiac valves.

## Conclusion

We present a validated rule-based NLP system that accurately extracts heart valve leaflet morphology from clinical echocardiographic reports. The system demonstrated excellent performance for BAV identification, enabling creation of the largest BAV patient cohort reported to date. This automated approach addresses the historical challenge of BAV identification in structured EHR data and facilitates large-scale retrospective cardiovascular research.

The successful implementation of NLP for cardiac valve assessment illustrates the broader potential for automated clinical text processing to enhance both research capabilities and clinical care. As healthcare systems increasingly recognize the value of unstructured clinical data, sophisticated NLP approaches will play an essential role in translating documented clinical observations into actionable insights for patient care and scientific discovery.

In this article, we present a custom NLP rule-based system that accurately extracts heart valve leaflet structure information from clinical notes. The initial use case for this data was the identification of BAV, which has not been available in structured data elements of the EHR until quite recently. The NLP system identified 84,019 patients with BAV from free-text echocardiography reports, making this the largest cohort of patients with BAV to our knowledge and enabling large-scale retrospective studies of this phenotype.

## Data Availability

Patient-level data are already accessible to all VA researchers with appropriate IRB approvals

## Abbreviations

BAV: bicuspid aortic valve
CDW: Corporate Data Warehouse
CPT: Current Procedural Terminology
EHR: electronic health record
ICD: International Classification of Diseases
NLP: natural language processing
VA: U.S. Department of Veterans Affairs

## Acknowledgements

This work was supported using resources and facilities of the Department of Veterans Affairs (VA) Informatics and Computing Infrastructure (VINCI), including NLP resources, which is funded under the research priority to Put VA Data to Work for Veterans (VA ORD 24-D4V-02).

This publication does not represent the views of the Department of Veterans Affairs or the United States Government.

