## Appendix for "Automated detection of bicuspid aortic valve from echocardiographic reports using natural language processing: a large-scale Veterans Affairs study"

The complete list of CPT codes used to select echocardiography procedures.

| Row ID | CPT Code | CPT Description |
| --- | --- | --- |
| 1 | 0613T | PERCUTANEOUS TRANSCATHETER IMPLANTATION OF INTERATRIAL SEPTAL SHUNT DEVICE, INCLUDING RIGHT AND LEFT HEART CATHETERIZATION, INTRACARDIAC ECHOCARDIOGRAPHY, AND IMAGING GUIDANCE BY THE PROCEDURALIST, WHEN PERFORMED |
| 2 | 0543T | TRANSAPICAL MITRAL VALVE REPAIR, INCLUDING TRANSTHORACIC ECHOCARDIOGRAPHY, WHEN PERFORMED, WITH PLACEMENT OF ARTIFICIAL CHORDAE TENDINEAE |
| 3 | 93315 | TRANSESOPHAGEAL ECHOCARDIOGRAPHY FOR CONGENITAL CARDIAC ANOMALIES; INCLUDING PROBE PLACEMENT, IMAGE ACQUISITION, INTERPRETATION AND REPORT |
| 4 | 0439T | MYOCARDIAL CONTRAST PERFUSION ECHOCARDIOGRAPHY, AT REST OR WITH STRESS, FOR ASSESSMENT OF MYOCARDIAL ISCHEMIA OR VIABILITY (LIST SEPARATELY IN ADDITION TO CODE FOR PRIMARY PROCEDURE) |
| 5 | 76828 | DOPPLER ECHOCARDIOGRAPHY, FETAL, PULSED WAVE AND/OR CONTINUOUS WAVE WITH SPECTRAL DISPLAY; FOLLOW-UP OR REPEAT STUDY |
| 6 | 93312 | ECHOCARDIOGRAPHY, TRANSESOPHAGEAL, REAL-TIME WITH IMAGE DOCUMENTATION (2D) (WITH OR WITHOUT M-MODE RECORDING); INCLUDING PROBE PLACEMENT, IMAGE ACQUISITION, INTERPRETATION AND REPORT |
| 7 | 3020F | LEFT VENTRICULAR FUNCTION (LVF) ASSESSMENT (EG, ECHOCARDIOGRAPHY, NUCLEAR TEST, OR VENTRICULOGRAPHY) DOCUMENTED IN THE MEDICAL RECORD (INCLUDES QUANTITATIVE OR QUALITATIVE ASSESSMENT RESULTS) (NMA-NO MEASURE ASSOCIATED) |
| 8 | 93351 | ECHOCARDIOGRAPHY, TRANSTHORACIC, REAL-TIME WITH IMAGE DOCUMENTATION (2D), INCLUDES M-MODE RECORDING, WHEN PERFORMED, DURING REST AND CARDIOVASCULAR STRESS TEST USING TREADMILL, BICYCLE EXERCISE AND/OR PHARMACOLOGICALLY INDUCED STRESS, WITH INTERPRETATION AND REPORT; INCLUDING PERFORMANCE OF CONTINUOUS ELECTROCARDIOGRAPHIC MONITORING, WITH SUPERVISION BY A PHYSICIAN OR OTHER QUALIFIED HEALTH CARE PROFESSIONAL |
| 9 | 3055F | LEFT VENTRICULAR EJECTION FRACTION (LVEF) LESS THAN OR EQUAL TO 35% (HF) |
| 10 | 76826 | ECHOCARDIOGRAPHY, FETAL, CARDIOVASCULAR SYSTEM, REAL TIME WITH IMAGE DOCUMENTATION (2D), WITH OR WITHOUT M-MODE RECORDING; FOLLOW-UP OR REPEAT STUDY |
| 11 | 93314 | ECHOCARDIOGRAPHY, TRANSESOPHAGEAL, REAL-TIME WITH IMAGE DOCUMENTATION (2D) (WITH OR WITHOUT M-MODE RECORDING); IMAGE ACQUISITION, INTERPRETATION AND REPORT ONLY |
| 12 | 3021F | LEFT VENTRICULAR EJECTION FRACTION (LVEF) LESS THAN 40% OR DOCUMENTATION OF MODERATELY OR SEVERELY DEPRESSED LEFT VENTRICULAR SYSTOLIC FUNCTION (CAD, HF) |
| 13 | 76825 | ECHOCARDIOGRAPHY, FETAL, CARDIOVASCULAR SYSTEM, REAL TIME WITH IMAGE DOCUMENTATION (2D), WITH OR WITHOUT M-MODE RECORDING; |
| 14 | 76827 | DOPPLER ECHOCARDIOGRAPHY, FETAL, PULSED WAVE AND/OR CONTINUOUS WAVE WITH SPECTRAL DISPLAY; COMPLETE |
| 15 | 93352 | USE OF ECHOCARDIOGRAPHIC CONTRAST AGENT DURING STRESS ECHOCARDIOGRAPHY (LIST SEPARATELY IN ADDITION TO CODE FOR PRIMARY PROCEDURE) |
| 16 | 93355 | ECHOCARDIOGRAPHY, TRANSESOPHAGEAL (TEE) FOR GUIDANCE OF A TRANSCATHETER INTRACARDIAC OR GREAT VESSEL(S) STRUCTURAL INTERVENTION(S) (EG, TAVR, TRANSCATHETER PULMONARY VALVE REPLACEMENT, MITRAL VALVE REPAIR, PARAVALVULAR REGURGITATION REPAIR, LEFT ATRIAL APPENDAGE OCCLUSION/CLOSURE, VENTRICULAR SEPTAL DEFECT CLOSURE) (PERI-AND INTRA-PROCEDURAL), REAL-TIME IMAGE ACQUISITION AND DOCUMENTATION, GUIDANCE WITH QUANTITATIVE MEASUREMENTS, PROBE MANIPULATION, INTERPRETATION, AND REPORT, INCLUDING DIAGNOSTIC TRANSESOPHAGEAL ECHOCARDIOGRAPHY AND, WHEN PERFORMED, ADMINISTRATION OF ULTRASOUND CONTRAST, DOPPLER, COLOR FLOW, AND 3D |
| 17 | 93303 | TRANSTHORACIC ECHOCARDIOGRAPHY FOR CONGENITAL CARDIAC ANOMALIES; COMPLETE |
| 18 | 93304 | TRANSTHORACIC ECHOCARDIOGRAPHY FOR CONGENITAL CARDIAC ANOMALIES; FOLLOW-UP OR LIMITED STUDY |
| 19 | 93356 | MYOCARDIAL STRAIN IMAGING USING SPECKLE TRACKING-DERIVED ASSESSMENT OF MYOCARDIAL MECHANICS (LIST SEPARATELY IN ADDITION TO CODES FOR ECHOCARDIOGRAPHY IMAGING) |
| 20 | 93307 | ECHOCARDIOGRAPHY, TRANSTHORACIC, REAL-TIME WITH IMAGE DOCUMENTATION (2D), INCLUDES M-MODE RECORDING, WHEN PERFORMED, COMPLETE, WITHOUT SPECTRAL OR COLOR DOPPLER ECHOCARDIOGRAPHY |
| 21 | 93306 | ECHOCARDIOGRAPHY, TRANSTHORACIC, REAL-TIME WITH IMAGE DOCUMENTATION (2D), INCLUDES M-MODE RECORDING, WHEN PERFORMED, COMPLETE, WITH SPECTRAL DOPPLER ECHOCARDIOGRAPHY, AND WITH COLOR FLOW DOPPLER ECHOCARDIOGRAPHY |
| 22 | 93317 | TRANSESOPHAGEAL ECHOCARDIOGRAPHY FOR CONGENITAL CARDIAC ANOMALIES; IMAGE ACQUISITION, INTERPRETATION AND REPORT ONLY |
| 23 | 93320 | DOPPLER ECHOCARDIOGRAPHY, PULSED WAVE AND/OR CONTINUOUS WAVE WITH SPECTRAL DISPLAY (LIST SEPARATELY IN ADDITION TO CODES FOR ECHOCARDIOGRAPHIC IMAGING); COMPLETE |
| 24 | 93325 | DOPPLER ECHOCARDIOGRAPHY COLOR FLOW VELOCITY MAPPING (LIST SEPARATELY IN ADDITION TO CODES FOR ECHOCARDIOGRAPHY) |
| 25 | 93350 | ECHOCARDIOGRAPHY, TRANSTHORACIC, REAL-TIME WITH IMAGE DOCUMENTATION (2D), INCLUDES M-MODE RECORDING, WHEN PERFORMED, DURING REST AND CARDIOVASCULAR STRESS TEST USING TREADMILL, BICYCLE EXERCISE AND/OR PHARMACOLOGICALLY INDUCED STRESS, WITH INTERPRETATION AND REPORT; |
| 26 | 93316 | TRANSESOPHAGEAL ECHOCARDIOGRAPHY FOR CONGENITAL CARDIAC ANOMALIES; PLACEMENT OF TRANSESOPHAGEAL PROBE ONLY |
| 27 | 93321 | DOPPLER ECHOCARDIOGRAPHY, PULSED WAVE AND/OR CONTINUOUS WAVE WITH SPECTRAL DISPLAY (LIST SEPARATELY IN ADDITION TO CODES FOR ECHOCARDIOGRAPHIC IMAGING); FOLLOW-UP OR LIMITED STUDY (LIST SEPARATELY IN ADDITION TO CODES FOR ECHOCARDIOGRAPHIC IMAGING) |
| 28 | 93662 | INTRACARDIAC ECHOCARDIOGRAPHY DURING THERAPEUTIC/DIAGNOSTIC INTERVENTION, INCLUDING IMAGING SUPERVISION AND INTERPRETATION (LIST SEPARATELY IN ADDITION TO CODE FOR PRIMARY PROCEDURE) |
| 29 | 93308 | ECHOCARDIOGRAPHY, TRANSTHORACIC, REAL-TIME WITH IMAGE DOCUMENTATION (2D), INCLUDES M-MODE RECORDING, WHEN PERFORMED, FOLLOW-UP OR LIMITED STUDY |
| 30 | 93313 | ECHOCARDIOGRAPHY, TRANSESOPHAGEAL, REAL-TIME WITH IMAGE DOCUMENTATION (2D) (WITH OR WITHOUT M-MODE RECORDING); PLACEMENT OF TRANSESOPHAGEAL PROBE ONLY |
| 31 | 93318 | ECHOCARDIOGRAPHY, TRANSESOPHAGEAL (TEE) FOR MONITORING PURPOSES, INCLUDING PROBE PLACEMENT, REAL TIME 2-DIMENSIONAL IMAGE ACQUISITION AND INTERPRETATION LEADING TO ONGOING (CONTINUOUS) ASSESSMENT OF (DYNAMICALLY CHANGING) CARDIAC PUMPING FUNCTION AND TO THERAPEUTIC MEASURES ON AN IMMEDIATE TIME BASIS |
| 32 | 93319 | NULL |

Keywords used to select reports:

‘”bicusp*” OR “bi-cuspid” OR “bi cuspid” OR “bileaflet” OR “bi-leaflet” OR “bi leaflet”’

### Annotation Guideline

**Overview**

We have been working on a project to identify the leaflet structures of the four valves of the heart. The echo reports you’ll be reviewing have been pre-annotated with a keyword dictionary, but we do not expect every term to have been captured. Create new annotations for any relevant concepts that the pre-annotator missed.

Annotation workflow:

1. Review the entire note and determine whether an echo report is present. Annotate the note title or the echo report section title and classify it as a primary echo report or other note type. If the document does not have an obvious title and there is no echo report within the document, highlight the first sentence and classify it with your best judgement.
2. If a heart valve and its corresponding leaflet structure was given and either of the terms were missed by the pre-annotator, annotate it.
   a. You do not have to highlight all heart valves in the document, only when the leaflet structure is present.
   b. You do not need to delete any pre-annotations. The annotations that are not connected by any relationships will be ignored.
3. Create relationships between the valve and its corresponding leaflet structure.
4. If there is a context phrase (uncertainty, negation, other experiencer, or historical), annotate it and create a relationship to the leaflet structure term.

The four valves of the heart:

1. Aortic valve: normally has a tricuspid leaflet structure.
   a. **We are specifically interested in identifying bicuspid AV.**
2. Mitral valve: normally has a bicuspid leaflet structure
3. Pulmonary valve: normally has a tricuspid leaflet structure.
4. Tricuspid valve: normally has a tricuspid leaflet structure.

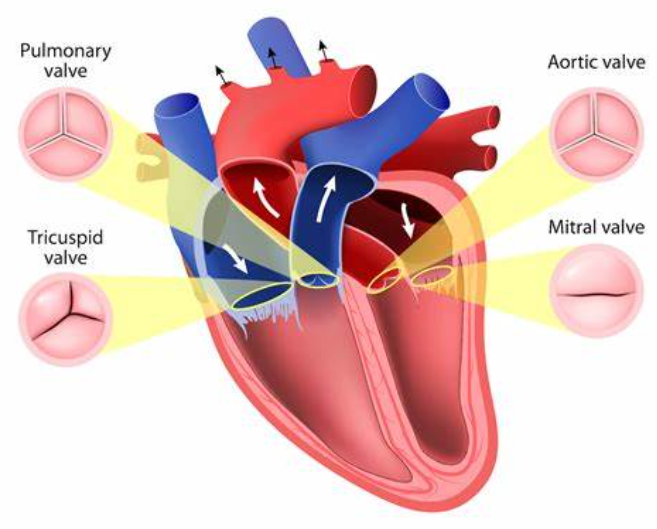

**Classes**

The annotation classes have been organized in the categories shown in the table below. Each class is described in further detail in the following sections.

| **Category** | **Class** |
| --- | --- |
| Note type | Document title |
| Valves | Aortic valve |
|  | Mitral valve |
|  | Tricuspid valve |
|  | Pulmonary valve |
|  | Prosthesis |
| Procedures | Valve replacement |
| Structures | Bicuspid structure |
|  | Tricuspid structure |
|  | Normal structure |
|  | Other structure |
| Context | Uncertainty |
|  | Negation |
|  | Other experiencer |
|  | Historical |

**Note Type Class**

This is a document-level annotation. The entire document may be a single echo report, or an echo report may be one section within a longer document. An echo report is likely to have a note title containing the word “echo”.

**Note type class 1: Document title**

Description: Review the note and determine the document title or echo report section header. If there is no obvious document title or echo report section header, highlight the first sentence. Select an attribute to classify the document, even if there is no obvious document title.

Attributes:

1. Primary echo report: the entire document is an echo report.
2. Primary echo report within note: if an echo report is a section within a longer document, highlight the echo section header as the document title, even if there is a document title for the overall document.
3. Other non-echo report type: valve-structure information is present, but the document does not contain an actual echo report.

Examples:

- Transthoracic echocardiography report
- Transthoracic echocardiogram
- Echo 8/10/2012
  - Can highlight the date of the procedure when it is right next to the echo term, but not necessary if the terms are separated by many tokens
- Artery and Vein Conditions (Vascular Diseases Including Varicose Veins) Disability Benefits Questionnaire

Do not annotate:

Relationships: N/A

**Valves Classes**

These are instance-level annotations, meaning there may be more than one per document. The four valves of the heart have been pre-annotated with some simple keywords, but we expect some terms to be missing. If a heart valve and its structure are present in the note, but were missed by the pre-annotator, manually highlight it. You do not have to annotate all heart valve terms in the document, only those where the leaflet structure is given.

**IMPORTANT:** Do not annotate the term “valve” by itself.

**Valve class 1. Aortic valve**

Description: The aortic valve of the heart. **This is the valve we are most interested in.** Normally tricuspid structure, but bicuspid in approximately ~1% of the population. Unicuspid and quadricuspid structures are even less frequent.

Examples:

- Aortic valve
- Aortic cusps
- AV
  - AV was not pre-annotated because “av” will appear in the middle of other words. Please annotate it with the “Aortic valve” class when you come across it.
- Aortic root and valve
- Aortic leaflets

Do not annotate:

- Aortic root
- Bicuspid valve
  - The mitral valve is normally bicuspid, so when clinicians use “bicuspid valve”, assume they are referring to the mitral valve. Only assume bicuspid aortic valve when “aortic” is explicitly used.
- AVR
  - This is an acronym for “aortic valve replacement”, so highlight it with the “valve replacement” class.

Relationships: Connect this to a structure class when present.

**Valve class 2. Mitral valve**

Description: The mitral valve of the heart. Normally bicuspid structure.

Examples:

- Mitral valve
- MV
- Bicuspid valve
  - The mitral valve is normally bicuspid, so when clinicians use “bicuspid valve”, assume they are referring to the mitral valve. Only assume bicuspid aortic valve when “aortic” is explicitly used.

Do not annotate:

- Bi-leaflet (St. Jude) mechanical prosthesis
  - If the mitral valve has been replaced with a prosthesis, annotate it using the “prosthesis” class described below.

Relationships: Connect this to a structure class when present.

**Valve class 3. Tricuspid valve**

Description: The tricuspid valve of the heart. Normally tricuspid structure. As shown in the screenshot below, the pre-annotator incorrectly captures some mentions of the tricuspid valve as “tricuspid structure”. Please check the class type and update if needed.

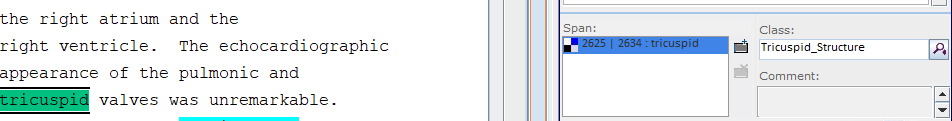

Examples:

- Tricuspid valve
- TV

Do not annotate:

- Tricuspid (by itself)
  - May need to use the surrounding note to disambiguate, but usually assume that when “tricuspid” is used by itself, it is referring to a structure type rather than the valve.

Relationships: Connect this to a structure class when present.

**Valve class 4. Pulmonary valve**

Description: The pulmonary valve of the heart. Normally tricuspid structure.

Examples:

- Pulmonary valve
- Pulmonic valve
- PV

Do not annotate: --

Relationships: Connect this to a structure class when present. Otherwise, no relationship.

**Valve class 5. Prosthesis valve**

Description: If a patient has a heart valve prosthesis, use this class to annotate the entire phrase. Delete any pre-annotations before creating this class. If both the valve and leaflet structure are given, highlight it all together as one annotation. Do not create a relationship between them.

Examples:

- Porcine aortic valve prosthesis
- Bi-leaflet (St. Jude) mechanical prosthesis

Do not annotate:

- Valve replacement in 2015
  - When the note describes the patient had a valve replacement procedure at some point, highlight it with the “valve replacement” class instead.

Relationships: N/A

**Valve class 6. Bicuspid AV generic statement**

Description: This is a new class designed to deal with generic statements about bicuspid aortic valve that are not referring to the patient. The pre-annotator will highlight bicuspid aortic valves from empty templates and clinical guidelines (see examples below). Delete the pre-annotations and highlight the valve and structure together with this class. You may include as many tokens as necessary to provide context.

Examples:

- [ ] BICUSPID AORTIC VALVE
- Literature reviewed: Egbe AC et al. Predictors of intracranial aneurysms in patients with bicuspid aortic valve.

Do not annotate:

- [X] BICUSPID AORTIC VALVE
  - If a template is filled in and it is clear the bicuspid aortic valve mention is affirmed for the patient, then create the valve-structure relationship as normal. Given the example above, a filled in checkbox can be considered affirmed. Do not highlight the [X], just the structure and valve terms.

Relationships: N/A – highlight the valve and structure terms together

**Procedure Class**

Note states that a patient has had a heart valve replaced at some point. Do not include the date of the procedure in the annotation. If the actual prosthesis type is detailed, use the “prosthesis” class instead. To differentiate between this class and the prosthesis class, use this class when a procedure (verb) is being referred to and use the prosthesis class when the actual prosthesis (noun) is detailed. You can also use this class when the word “replacement” is used.

**Procedure class 1. Valve replacement**

Description: When the note describes the patient had a valve replacement procedure at some point, highlight it with the “valve replacement” class. This can be a replacement of any of the four heart valves. Delete any pre-annotations before creating this class. If both the valve and leaflet structure are given, highlight it all together as one annotation and do not create a relationship.

Examples:

- AVR
  - Acronym for “aortic valve replacement”.
- Valve replacement in 2015
- Pt with recent bioprosthetic aortic valve replacement, CABG and CVA s/p surgery.
- Patient with known HTN, AS s/p porcine AVR—needs routine eval of valve replacement
  - Highlight both “porcine AVR” and “valve replacement” as valve replacement
- Indication: CAD s/p CABG; Mitral and aortic valve replacement
  - If multiple valves are given in one span, highlight as one.
- Hx coarctation of aorta, aortic stenosis and bicuspid aortic valve with aortic valve replacement (Freestyle 29mm valve) and aortic repair 04/2009.
  - Highlight “bicuspid” as bicuspid structure, “aortic valve” as aortic valve, “aortic valve replacement” as valve replacement, and “Freestyle 29mm valve” as prosthesis valve.
  - Create a relationship between bicuspid and aortic valve.

Do not annotate:

- Recommend cardiology consultation for AV replacement.
  - The patient has not actually had a replacement yet, so do not highlight it.

Relationships: There are no relationships for this class. Even if context is given (usually historical), you do not need to highlight and attach it to this class.

**Structure Classes**

For this project, structure refers to the leaflet structure of the heart valve. Structure terms include “tricuspid”, “trileaflet”, “bicuspid”, “bileaflet” or “normal”.

**Structure class 1. Tricuspid structure**

Description: The heart valve leaflets have a tricuspid structure. May need to use the surrounding note to disambiguate, but usually assume “tricuspid” is referring to the leaflet structure type rather than the actual valve.

Examples:

- Trileaflet
- “The aortic valve is normal with a tricuspid structure.” – When normal and tricuspid are in the same sentence, highlight them together and use the “Tricuspid Structure” class.
- “The aortic valve is normal. The valve is tricuspid.” – In this instance, normal and tricuspid are in separate sentences. Most NLP programs work on the sentence level, so the structure term in the same sentence as the specific valve term has the highest priority. Connect “aortic valve” to “normal”, not “tricuspid”. “Tricuspid” will be pre-highlighted by the pre-annotator; you do not have to delete it. Do not highlight the second “valve” that appears by itself.
-
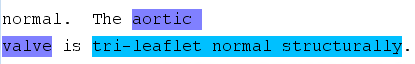

  - Highlight “tri-leaflet” and “normal structurally” all together as “tricuspid structure”.

Do not annotate:

- Normal (by itself)
  - If the valve is normally tricuspid and is described as “normal” (the word tricuspid is not used in the note), use the “Normal Structure” class (described below), not the “Tricuspid Structure” class.

Relationships: Connect this to a valve class.

**Structure class 2. Bicuspid structure**

Description: The valve has a bicuspid structure. Classify the bicuspid type as congenital, functional, or unspecified (default) with the attribute. Highlight “congenital” or “functional” with “bicuspid” if they are close together, but it is not necessary if they are far apart. If no specific terms are present, leave as “unspecified” (default). This is most important for bicuspid aortic valve, but if other valves are indicated to be congenitally or functionally bicuspid, you can change the attribute.

Attributes:

1. Congenital: specifically stated that the bicuspid aortic valve is congenital.
   a. Example: “*Congenital bicuspid aortic valve.”*
2. Functional: specifically stated that the valve is functionally bicuspid
   a. Example: “*Aortic valve: The valve was likely functionally bicuspid and had fusion of the right-noncoronary commissure.*”
3. Unspecified (default): there are no specific terms to indicate whether the valve is functionally or congenitally bicuspid.
   a. Example: “*Aortic valve is heavily calcified. There appears to be a partial fusion of the left and non-coronary cusp, giving a bicuspid appearance to the valve.”*
      i. Specific congenital and functional terms are not present, so classify as “unspecified”.

Examples:

- Bicuspid
- Bileaflet
-
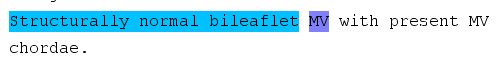

  - Another example where normal and bileaflet are in the same sentence and should be highlighted together as “Bicuspid structure”.
-
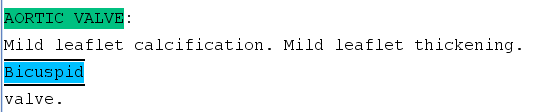

  - Do not annotate “bicuspid valve”, only “bicuspid”.

Do not annotate:

- Normal structure
  - If the valve is normally bicuspid and it is stated that the valve is normal (with no other more specific structure term), use the Normal Structure class (described next), not the Bicuspid Structure class.
- “Hx aortic stenosis with bicuspid aortic valve replacement 2008 and aortic stent done at Duke.”
  - Delete pre-annotations “bicuspid” and “aortic valve”, highlight “bicuspid aortic valve replacement” as “valve replacement”.

Relationships: Connect this to a valve class.

**Structure class 3. Normal structure**

Description: The valve is described as having a normal structure and there is no other, more specific, structure term present.

Examples:

- Normal
- Normal structure
- Structurally normal
- Anatomically normal
- Unremarkable

Do not annotate:

- “The aortic valve has a normal, tricuspid structure.”
  - Annotate “normal, tricuspid structure” with the Tricuspid Structure class, not the Normal Structure class.
- “The tricuspid valve opens normally.”
  - As seen in the example below, it is stated that the tricuspid, mitral, and aortic valve all open normally. Since it is stated separately that the aortic valve is possibly bicuspid, do not annotate “opens normally” as Normal Structure for any of the valves.

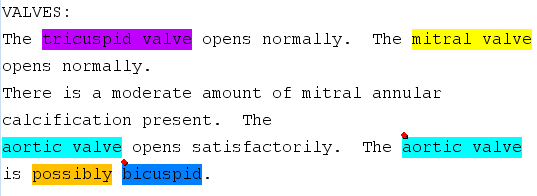

Relationships: Connect this to a valve class.

**Structure class 4. Other structure**

Description: Only use this class for unicuspid and quadricuspid valve structures. These abnormalities are extremely rare, but if these structures are described in the notes, mark them as “Other structure” and create a relationship to the corresponding valve.

Examples:

- Quadricuspid
- Unicuspid

Do not annotate: --

Relationships: Connect this to a valve class.

**Context Classes**

**Context Class 1: Uncertainty**

Description: Highlight terms/phrases indicating uncertainty about a valve structure. Create a relationship between the uncertainty phrase and the structure term.

Examples:

- Bicuspid AV cannot be totally excluded.
- Cannot rule out bicuspid aortic valve.
- Looks like it could be a bicuspid aortic valve.
- Probably bicuspid aortic valve. However this cannot be confirmed with the limited views.
  - Do not need to annotate anything in the second sentence. Only highlight “probably”.
- Consider further evaluation for bicuspid aortic valve and need for replacement if necessary.
  - “Further evaluation” can be considered an uncertainty modifier of bicuspid aortic valve. Do not highlight “replacement”, as the patient has not yet had a valve replacement.

Do not annotate:

- “The aortic valve appears to be a tricuspid structure.”
  - We will assume “appears to be” to be a positive assertion, not uncertain.

Relationships: Connect uncertainty to the structure class, not the valve.

**Context Class 2: Negation**

Description: Use this class to highlight terms/phrases that indicate a valve structure is negated.

Examples:

- The aortic valve does not have bicuspid structure.
- Bicuspid aortic valve: No

Do not annotate: --

Relationships: Connect negation to the structure class, not the valve.

**Context Class 3: Other experiencer**

Description: Use this class to highlight terms/phrases that indicate a valve structure is referring to an experiencer other than the patient (usually a family member).

Examples:

- The patient’s brother and father both have bicuspid aortic valves.

Do not annotate: --

Relationships: Connect other experiencer to the structure class, not the valve.

**Context Class 4: Historical**

Description: Use this class to highlight terms/phrases that indicate a valve structure is a historical mention. If a valve replacement procedure is modified with a historical statement, highlight the valve replacement class, but not the historical statement. We do not care about context related to valve replacements, only valve structures. If there is no explicit term to indicate history, do not highlight anything (see do not annotate example).

Examples:

- H/o bicuspid aortic valve.

Do not annotate:

- Valve replacement in 2015 for congenital bicuspid aortic valve.
  - Humans can understand that the patient had a bicuspid aortic valve in the past, but there is no distinct term that can be highlighted to tell this to the computer. The goal of this project is to identify all patients who had a bicuspid aortic valve at any point, so it is okay that we can’t flag this as a historical mention. Highlight “valve replacement” with the valve replacement class and create a relationship between the “bicuspid” and “aortic valve” classes.
- H/o valve replacements
  - Only highlight “valve replacements”, not “h/o” because we do not care about context related to valve replacement procedures.

Relationships: Connect historical to the structure class, not the valve class.

**Relationships**

To create relationships, click on the plus sign next to the Relationships section of the Annotation Editor side bar. When it shows a pen drawing, you can create relationships by left clicking on Class 1 (see table below) and right clicking on Class 2.

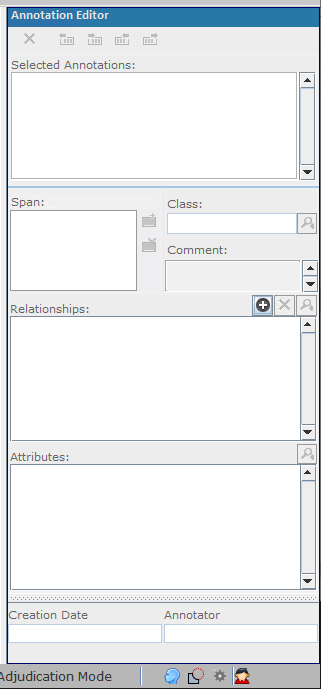
 🡪
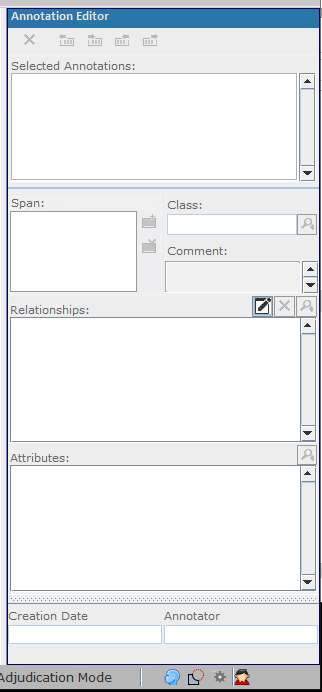

Create relationships between the heart valve and its corresponding structure term, when present. Most NLP programs work on the sentence level, so when creating relationships between the valve and structure terms, give the highest priority to the structure term in the same sentence as the valve term. If context is present in the note, create a relationship between the structure term and the context term.

| **Class 1** | **Class 2** | **Description** |
| --- | --- | --- |
| [Aortic \| Mitral \| Pulmonary \| Tricuspid] valve | [Bicuspid \| Tricuspid \| Normal \| Other] structure | Connect a valve term to a structure term, when present. If “valve” by itself is closest to the structure term, connect the structure term to the closest *specific* valve term. **A single structure term can be connected to multiple valve terms, but do not connect more than one structure term to a valve term (see example in the next section.)** |
| [Bicuspid \| Tricuspid \| Normal \| Other] structure | [Uncertainty \| Negation \| Other experiencer \| Historical] context | If context is present in the note, create a relationship between the structure term and the context term. Do not connect the context term to the valve. |

**Additional Examples**

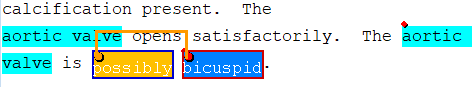

- When uncertainty or negation is present, create a relationship between the uncertainty/negation term and the leaflet structure term. Do not connect it to the valve term.

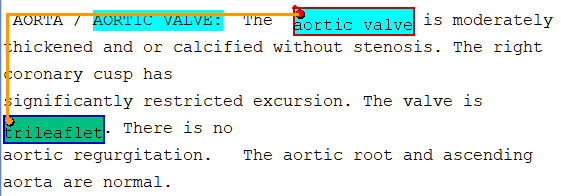

- Do not highlight the term “valve” by itself. Connect the structure to the closest, specific valve term.
  - In this example, both aortic valve terms were highlighted by the pre-annotator. Normally we prefer to have the valve term and structure term in the same sentence. In this case, the sentence with the structure term only uses the word “valve”, which is not specific enough, so the structure term is connected to the closest aortic valve term. If cases the closest specific valve term is the section header, the structure term can be connected to the section header.

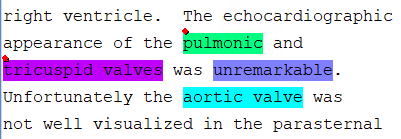

- Do not highlight “valve”, but “pulmonic”, “mitral”, etc. can be highlighted individually.
  - Highlight “unremarkable” as “Normal structure”. Connect both “pulmonic” and “tricuspid valves” to “unremarkable”. When it states that a valve was not well-visualized, as with the aortic valve here, there is no indication on the type of leaflet structure, so do not highlight anything or create any relationships.

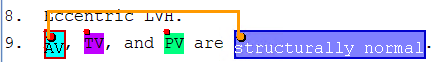

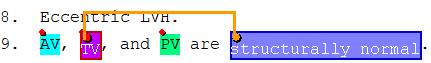

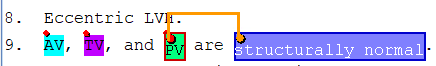

- A single structure term can be connected to multiple valve terms, but do not connect more than one structure term to a valve term.
- In this example, AV, TV, and PV are all connected to “structurally normal”.

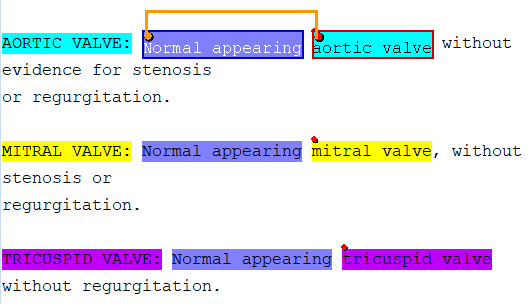

- When a specific valve term is given within the section, create a relationship between the valves and structures within the section, not the section headers. Section headers will have been pre-annotated by the keyword search.
  - UPDATE: in the example above, do not highlight “appearing”, only “normal”.

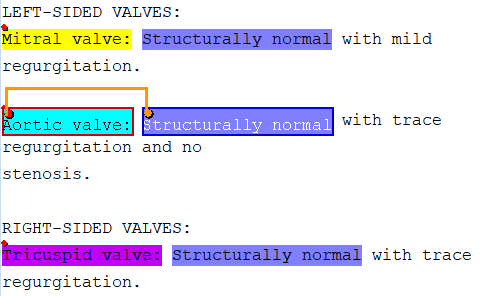

- When the section header is the closest, most specific valve term, create a relationship between the section header and the leaflet structure term.

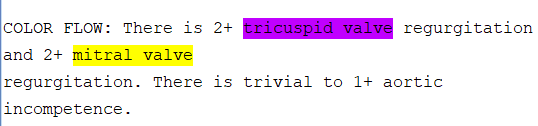

- Valves have been highlighted by the pre-annotator, but the leaflet structures are not given in the note. No additional classes to highlight or relationships to create.
